# A Metadata-Driven Framework for Strengthening Pathogen Genomics Lessons from SARS-CoV-2

**DOI:** 10.1101/2025.11.04.25339514

**Authors:** Michael J. Pavia, Karen O’Connor, Graciela Gonzalez-Hernandez, Matthew Scotch

## Abstract

During the COVID-19 pandemic, large-scale pathogen sequencing generated millions of SARS-CoV-2 genomes deposited in repositories like GenBank and GISAID. However, most of these records lack detailed patient metadata, such as demographics and clinical outcomes, which limits their utility for large-scale pathogen genomics analyses. While records that are linked to a journal publication might contain such metadata, systematic extraction and linkage to sequence records requires substantial manual effort. In this work, we assess the completeness of metadata in GenBank and demonstrate the value of enriched clinical and demographic annotations for genomic epidemiology. We found that on average GenBank records contained only 21.6% of host metadata, and during our study period ∼0.02% of published articles provided accessible sequence-specific patient metadata. Additionally, using published SARS-CoV-2 genomes and their corresponding journal articles, we constructed an analytical use case in pathogen genomics in which host stratification by clinical and demographic factors enables examination of evolutionary dynamics and clinical outcomes. Our results demonstrate how metadata-enrichment enhances pathogen genomic studies and provide a framework applicable to other pathogens.

## Introduction

The COVID-19 pandemic brought about significant changes in pathogen genomics, with massive amounts of viral sequences being shared openly in nucleotide repositories like GenBank [1] and GISAID [2]. These databases are essential for answering scientific questions on SARS-CoV-2 or other pathogens including differences in rates of evolution [3], transmission patterns [4], and geographic differences in diversity of variants of concern (VOC) [5]. While these platforms facilitate rapid sharing of genomic data, their utility is limited by inconsistent or incomplete reporting of patient metadata such as demographics, clinical outcomes, and/or comorbidities [6,7]. This metadata may be described in the accompanying publications, yet sequence records often lack a manuscript reference, either because one does not exist, or because a record was never updated to include the publication link [8]. Consequently, the lack of metadata in pathogen genomics limits our capacity to connect viral sequences with patient phenotypes, impeding the identification of key epidemiological patterns.

Integrating metadata with viral genomic data is essential for understanding how viral variation contributes to clinical outcomes. For example, we know that genetic variations in SARS-CoV-2 spike glycoprotein and host factors, such as ACE2 and TMPRSS2 polymorphisms, directly influence viral infectivity and disease severity [9]. In addition, while comparative analyses across variants have demonstrated that hospitalization rates vary by lineage [10], the associations often lose statistical significance once models account for population-level factors such as changes in clinical care standards and testing practices [11]. Further, without comorbidity data such as obesity (as measured by BMI), the impact of the A20268G mutation on hospitalization would have been attributed only to viral lineage [12].

Understanding the relationship between viral genotypes and patient clinical outcomes is dependent on the granularity of available metadata. For instance, *Patel et al.* [13] linked SARS-CoV-2 mutation profiles to age and geography to find that working age individuals (18-64 years) carry the highest burden of unique mutations, with high regional variability. Incorporating vaccination phases metadata further showed distinct spikes in unique mutations that coincided with vaccine rollout phases, indicating that demographics, geography, and public health initiatives together influence evolutionary rates. In another example, longitudinal sequencing of SARS-CoV-2 infections in immunosuppressed patients receiving antiviral treatments showed that these individuals harbor a greater number of private (non-lineage) mutations, which would have been missed in coarser datasets [14]. Furthermore, Larsen *et al.* [15] found that the spike protein mutation D614G is linked to shifts in symptom progression (a tendency for cough to precede fever); a relationship emerging only after the inclusion of detailed clinical symptom data into the analysis. These studies demonstrate that integrating detailed, high-quality patient metadata is critical for clarifying the clinical and public health implications of viral evolutionary dynamics.

Incomplete or ambiguous metadata poses significant challenges for accurate genomic epidemiology. As an example, in a global analysis of SARS-CoV-2 sequences from GISAID, it was observed that 63% of records lacked demographic data and more than 95% were missing patient-level clinical information [16]. Additionally, we have described the lack of host location metadata in pathogen sequence records in GenBank [17,18]. To address this issue, we have developed a framework integrating uncertainty in sampling locations into phylodynamic analysis, rather than relying on fixed geographic assignments [19,20]. This approach outperforms conventional methods when reconstructing viral persistence time, migration rates, and ancestral origins. Additionally, efforts have been made to develop a natural language processing (NLP) based system designed to automatically extract and refine geospatial data directly from scientific literature [21].

The goal of this study is to demonstrate how integrating host metadata with pathogen sequence data can improve our understanding of infectious disease outbreaks, using SARS-CoV-2 as the primary case study. We present an investigative analysis focusing on how host characteristics influence viral evolutionary dynamics and their clinical relevance. By leveraging metadata-enriched pathogen genomes we demonstrate an untapped potential for informing more effective responses to emerging viral threats.

## Methods

### Systematic search, screening, and data enrichment

We searched LitCovid [22] for full-text PubMed Central (PMC) articles published between January 2023 and December 2024. We used regular expressions to screen for (1) mentions of sequence databases (GenBank, BioProject, BioSample, SRA, and GISAID), (2) strings that matched the alphanumeric GenBank accession number format, and (3) references to variants of interest (VOI) or variants under monitoring (VUM) including BA.2, BA.2.86, JN.1, KP.2, KP.3, KP.3.1.1, LB.1, XEC, JN.1.7, and JN.1.18. Two reviewers independently assessed articles for all three mentions. We specified an inclusion criterion where an article needed to report (1) original SARS-CoV-2 complete genome sequence data, (2) the deposit of the raw reads or consensus sequences to GenBank, and (3) sequence-specific patient metadata derived from human hosts. For this effort, we only considered sequences deposited in GenBank in order to leverage the NCBI *Entrez* environment which links PMC and GenBank (nucleotide) databases [23]. We therefore excluded articles that submitted to GISAID or were derived from non-human hosts or from environmental sources such as wastewater.

For articles that met our inclusion criteria, we manually extracted sequence-specific patient metadata encompassing sample collection details, patient demographics, treatment regimens, laboratory results, vaccination status, infection presentation, and clinical outcomes when available. We defined enriched metadata as the patient information that was not reported in GenBank, whereas pre-enriched refers to the metadata that was recovered from Genbak, this metric was different for each article. Additionally, for each article where enriched metadata contained enough demographic and clinical information, such as age and comorbidities, we calculated the Charlson Comorbidity Index (CCI) [24]. To ensure consistency across studies, we grouped synonymous terms (including sampling location/method, comorbidities, and treatment coding) extracted from articles according to SNOMED CT [25]. We did not group ambiguous terms but rather left them as initially extracted.

### Sars-CoV-2 genome retrieval and assembly

Using the NCBI Entrez Direct [26] we obtained SARS-CoV-2 genomes from GenBank and SRA libraries for eight of the studies. We assembled the raw Illumina reads by first removing low quality reads and adapters with fastp [27], followed by IRMA for reference-based assembly [28]. For three studies where both GISAID genomes and SRA libraries were available, we compared our assembled genomes to those in GISAID using BLAST [29], we applied a threshold of >99.5% identity similarity or fewer than six combined mismatches and gaps; assemblies that did not meet these criteria were excluded from downstream analysis. We used Nextclade [30] to assign clades, evaluate the genome quality, and identify both nucleotide and amino acid mutations relative to Wuhan-1 genome [NC_045512.2]. Finally, we excluded from downstream analysis any genome classified as “bad” by Nextclade [30] for their overall QC status.

### Sequence selection and metadata integration for within-host analyses

While genomic data alone can be used to identify amino acid mutations, linking or stratifying these observations with clinical outcomes requires integration with patient metadata. Using the enriched dataset, we studied how the addition of immune status, comorbidities, and treatment regimen metadata improve our ability to understand viral evolution and patient outcomes for SARS-CoV-2 infections. To strengthen generalizability, we combined data from multiple articles where longitudinal SARS-CoV-2 sequence data was available as well as corresponding patient metadata encompassing immune status, treatments, and outcomes. This curated dataset consisted of 100 genomes from 34 patients (Figure 1, Table S1): Gonzalez_Reiche_2023 [31], Ignari_2024 [32], Manuto_2024 [33], and Pavia_2024 [34].

**Figure 1.**
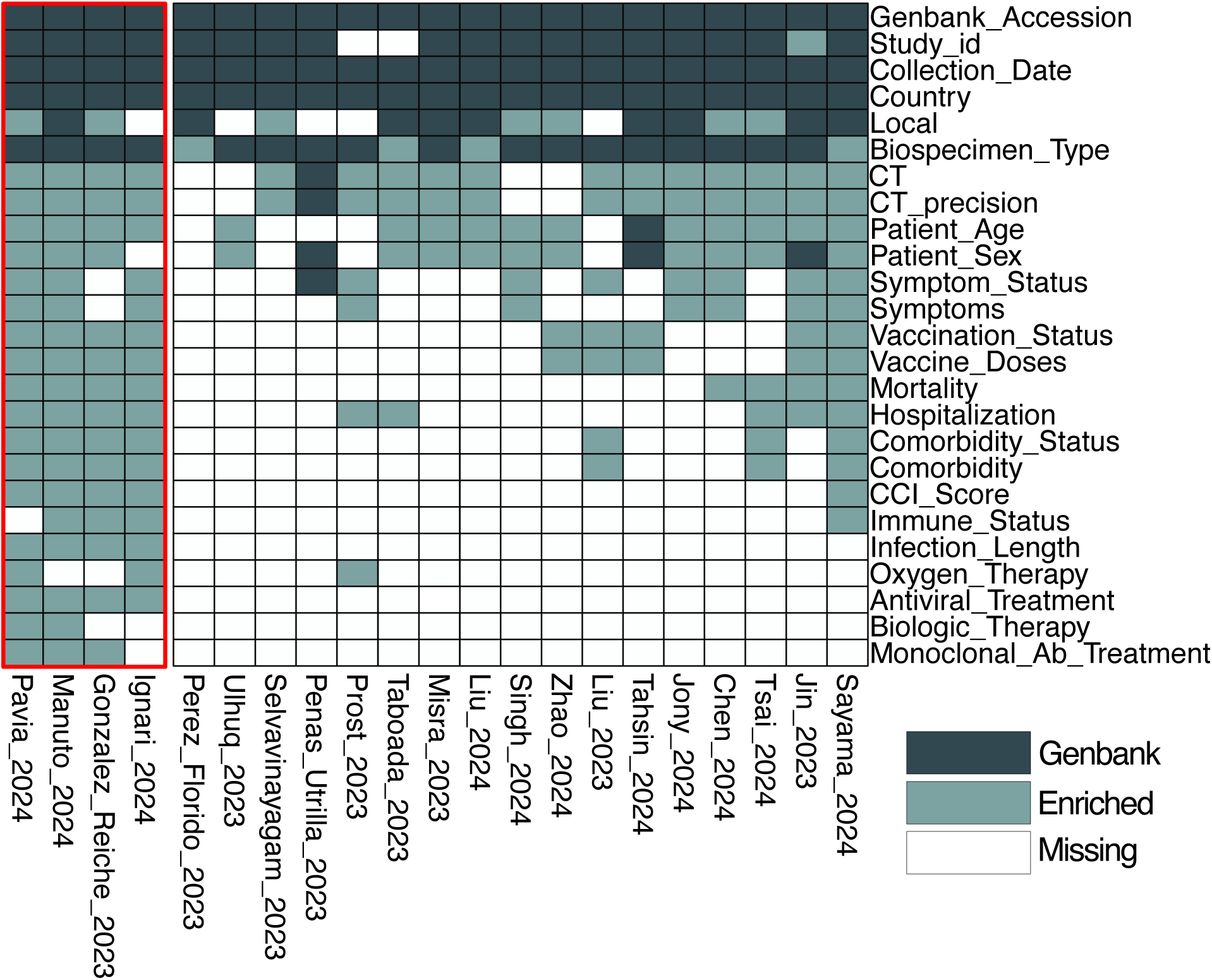
Heatmap of sequence-specific patient metadata collected. Dark blocks represent metadata in GenBank, light blocks represent metadata extracted from publication text, and white blocks represent missing metadata. Studies highlighted in red were used for in the case study. Local refers to state or city versus county level location. CT refers to RT-PCR cycle threshold with CT_precision indicating either exact value or if a range was reported. Symptom_status refers to either asymptomatic or symptomatic cases and Symptoms lists the reported symptoms. Vaccination_Status refers to whether a patient was vaccinated and Vaccine_dose specifies the number of doses received prior to sequencing. Comorbidity_Status refers to the presence or absence of comorbidities and Comorbidity lists the specific conditions reported.

### Case study: linking immune status to within-host evolution and outcomes via enriched metadata

We analyzed longitudinal SARS-CoV-2 genomes from four articles encompassing individuals with varying immune status, comorbidities, and treatment regimens. We performed phylogenetic reconstruction using IQ-TREE (GTR + G model) [35] with 1,000 bootstrap replicates. For time calibration, we used the treedater package [36] in R under an uncorrelated clock model. We estimated evolutionary rates for patients with two or more timepoints by fitting a linear regression to the root-to-tip genetic distance against the sampling dates and further assessed the dependence of these rates (mutations per site per year per patient) on immune status and viral clade with a multiple linear regression. We identified non-random recurrent mutations by applying an empirical binomial model to estimate the expected frequencies of amino acid and nucleotide mutations across the cohort. The resulting binomial p-values were then adjusted using the Benjamini-Hochberg method [37]. We defined empirical recurrence thresholds as the minimum number of patients for which the mutation’s observed frequency yielded a cumulative binomial probability < 0.05. Finally, we classified as *recurrent* any mutations that exceeded the threshold and appeared in ⋧2 viral clades. We set recurrence thresholds at a minimum of 11 patients for amino acid mutations and 10 patients for nucleotide mutations (Figure S1).

To assess associations between amino acid mutations, patient risk factors (immune status and CCI), treatment regimens, and outcomes (mortality, hospitalization, and infection length), we used generalized linear models with logistic regression for binary outcomes (mortality and hospitalization) and linear regression for infection duration. We grouped recurrent amino acid mutations with identical patient-level patterns to prevent redundancy and reduce model complexity. Our models consisted of: (1) mutation group only, (2) mutation group plus antiviral treatment, (3) mutation group plus patient factors, and (4) mutation group with antiviral treatment and patient factors. To guard against overfitting, we performed 5-fold cross validation [38]. We assessed the fit of the model to the data using ΔAIC and ΔBIC which we calculated relative to Δ = 0. We performed all statistical analyses in R v.4.4.2 and considered results with p-values < 0.05 as statistically significant.

## Results

### Availability of patient metadata in GenBank

A systematic screening of 116,600 recent publications from LitCovid identified a final set of 21 articles that met all inclusion criteria for metadata enrichment (Figure S2). In 2023, a total of 71,692 publications were listed in LitCovid, of which 68% (n = 48, 959) were open access and eligible for further analysis. In contrast, in 2024 there were only 44,908 publications with 49% available (n = 21,790) as open access. After applying regular expressions to identify GenBank accessions, sequence databases, and SARS-CoV-2 variants, we considered 640 and 442 candidate articles in 2023 and 2024, respectively. Through manual review, we narrowed the list to 21 total articles that met all inclusion criteria (10 in 2023 and 11 in 2024) and subjected these to comprehensive metadata collection (Table S1).

The studies included in our analysis predominantly sampled SARS-CoV-2 during 2022 (76.2%), coinciding with the emergence and global dominance of the Omicron variant and its sublineages [39]. Geographically, most studies were conducted in Asia (61.9%), followed by Europe (28.6%) and the Americas (9.5%). Across the 21 included articles, only sample collection date and country of origin were consistently reported in GenBank (Figure 1). In contrast, 14 clinically relevant metadata types were solely available in the article text and required manual enrichment.

We found a considerable discrepancy between the metadata reported in GenBank records alone and metadata available in the text of the article (and supplementary material). Across all studies, there was a wide range of metadata completeness, measured as the number of extractable metadata from just our articles. GenBank records captured only an average of 21.6% of host metadata across studies leaving more than 75% missing. Manual enrichment did improve overall completeness by an average of 30% recovering an additional 56% of metadata types absent from GenBank. Demographic variables such as age (71.4% of articles) and sex (61.9%) were primarily recovered through enrichment. However, disease outcomes (hospitalization, mortality, and symptoms) were reported less frequently: mortality was found in 38.1% of articles, hospitalization in 42.9%, and symptoms in 47.6%. Reporting was the lowest for infection length (19% of articles) and specific treatment regimens, with frequencies for oxygen therapy, antivirals, biologics, and monoclonal antibodies ranging from 9.5% to 14%.

Some studies, such as Manuto_2024 (37 genomes) and Pavia_2024 (9 genomes), reported nearly all metadata types extractable in our dataset (96% complete), representing best practices for data sharing and facilitating downstream analysis or cross-cohort comparisons. In contrast, Penas_Utrilla_2023 (6 genomes) baseline GenBank metadata was not improved by manual enrichment.

### Evolutionary dynamics and risk predictors in persistent SARS-CoV-2 infections

Nextclade placed our 100 sequences (34 patients; four articles) into five distinct PANGO Lineages, led by BA.1 and BA.5, with contributions from Italy, Japan, and the USA (Figure S3). Excluding collection date, this represents the analytical limits of metadata deposited in GenBank for these sequences. We conducted a multiple linear regression analysis to model the evolutionary rate (mutations per site per year per patient) as a function of both immune status and viral clade (Figure 2A). We show that immunocompromised individuals were significantly associated with higher within-host evolutionary rates (p = 0.016). Among clades, BA.1 (21K) had a near-significant association with higher rates of amino acid mutations (p=0.051), while the other clades showed no significant differences. We tested whether the diversity of unique mutations differed between immunocompromised and immunocompetent patients at nucleotide and amino acid levels, motivated by the effect of immune status on selection, infection duration, and thus the effective evolutionary substitution rate of the pathogen (Figure 2B). While the total number of nucleotide changes did not differ significantly between the two groups, immunocompromised individuals did harbor a significantly higher number of unique amino acid mutations (p=0.044). A total of 497 unique nucleotide mutations were identified (299 missense, 143 synonymous, 38 intergenic, and 17 deletions), with these resulting in 348 unique amino acid mutations.

**Figure 2.**
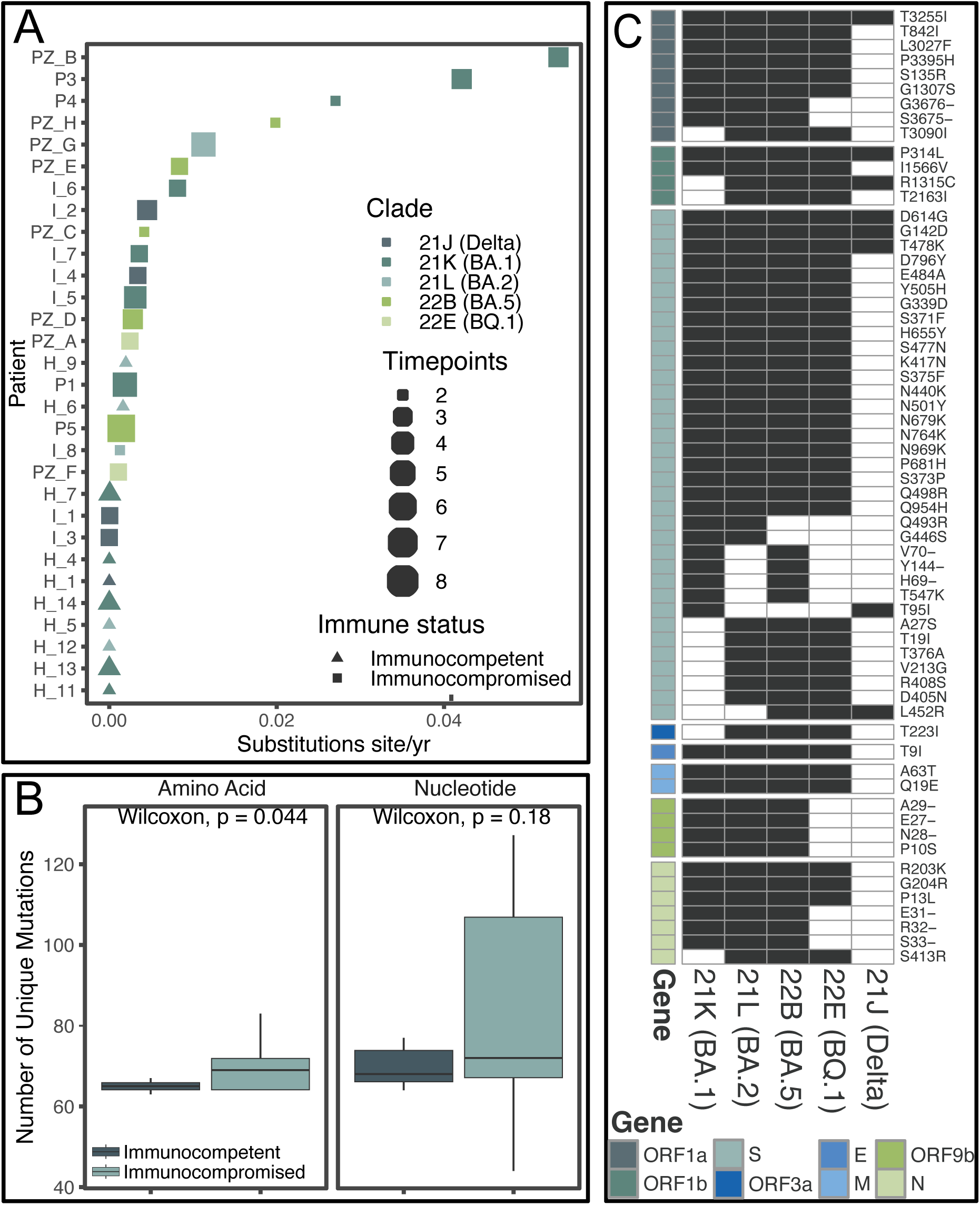
Mutation burden and recurrence across immune states and viral clades. (A) Dot plot of mutation rate per patient. (B) Boxplots comparing the number of unique amino acid and nucleotide mutations between immunocompetent and immunocompromised patients, p-values from Wilcoxon tests are shown. (C) Heatmap of recurrent amino acid mutations by clade. Black cells represent mutations in each clade (column), white cells represent their absence, and the adjacent color code indicates the corresponding gene for each mutation.

To categorize recurrent mutations from those arising by chance, we performed a rank-frequency analysis and used an empirical binomial model to set the patient-count threshold for which a mutation is unlikely to be a chance event (Figure S1). Applying this threshold, we identified 63 recurrent amino acid changes and 69 synonymous nucleotide mutations (Figure 3C). Among the recurrent mutations we identified two nucleocapsid mutations R203K + G204R and the Δ31-33 deletion, both of which are hallmark mutations of the Omicron lineage [40].

**Figure 3.**
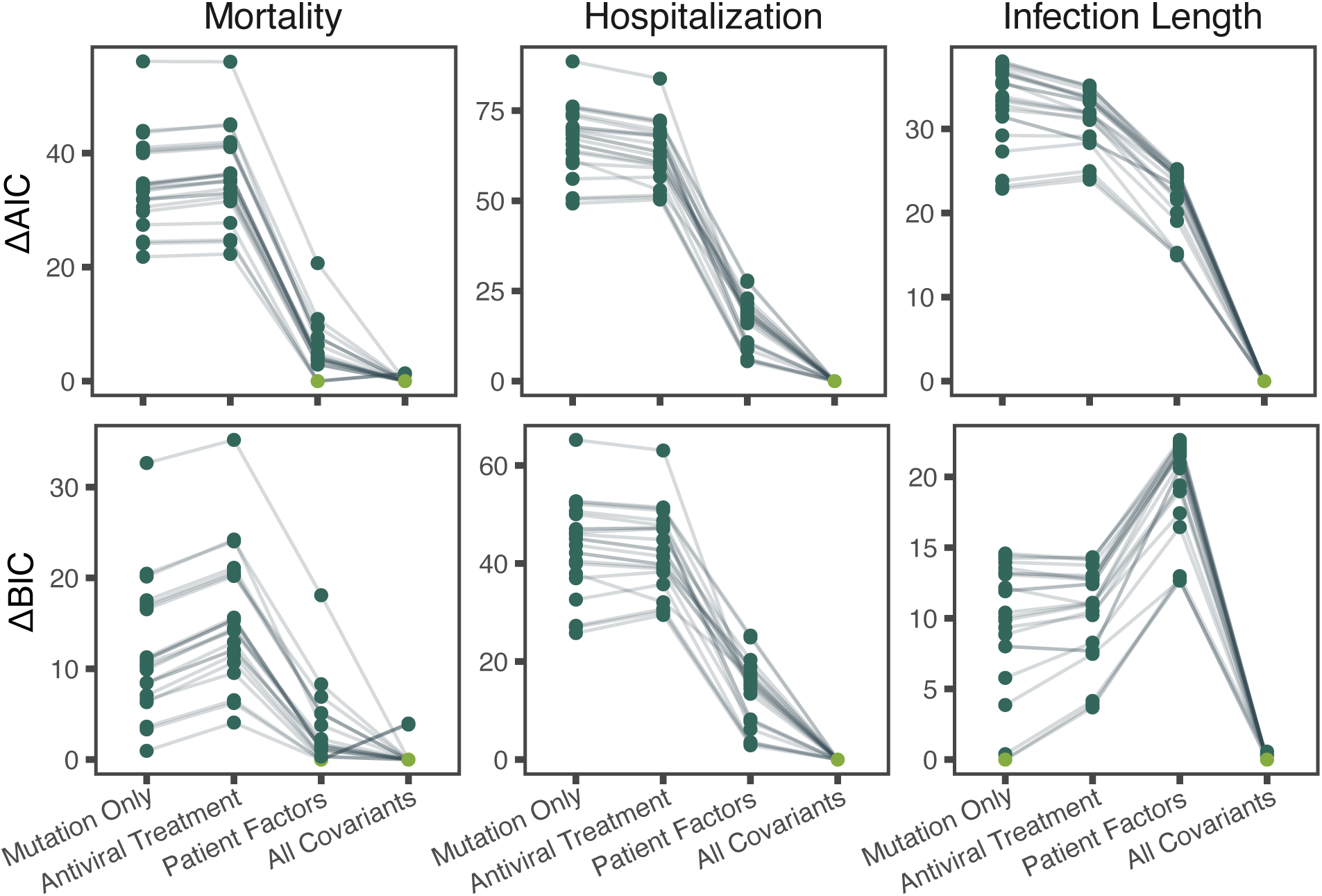
Model comparison for clinical outcomes using ΔAIC and ΔBIC. Vertical faceted plots represent the outcome tested by each model. Patient factors include age, CCI, and immune status. Lines link mutation groups from model to model. The best-fitting model (lowest ΔAIC or ΔBIC) per-mutation is highlighted in green.

Additionally, we identified two rare mutations, I1566V (ORF1b) and P10S (ORF9b), present in less than 0.01% of global sequences [41]. Every recurrent nucleotide mutation corresponded to an amino acid change, except for two intergenic mutations, C241T and A28271T. Notably, 35 of these recurrent amino acid mutations (55.5%) were localized to the spike protein, D405N, E484A, G446S, K417N, L452R, N440K, N501Y, N969K, Q493R, and S371F We contextualized the amino acid mutations by first grouping them based on congruent patient level presence and by integrating these groups with patient metadata, including immune status, CCI, and treatment regimens. To evaluate how the addition of clinical and treatment metadata improves the prediction of patient outcomes, we fit a series of nested regression models from mutation-only (equivalent to pre-enrichment analytical capabilities) to those fully adjusted for treatment regimen and patient characteristics (Figure 3). Across 23 groups (73.9% singletons), the fully adjusted models (including all covariates) demonstrated the strongest performance for ΔAIC and ΔBIC scores in 82.6% of groups for mortality, 100% of groups for hospitalization, and 95.7% of groups for infection duration.

We found two mutation groups and ten singletons to be significantly associated with the tested clinical outcomes (Table 1). For mortality, two groups showed statistical significance: notably a large cluster including structural genes (mutations in S and N) and non-structural genes (mutations in ORF1ab and ORF3a). The second group showing significant association with mortality was a collection of four mutations all occurring within ORF1a. Significant, amino acid mutations associated with hospitalization were primarily localized to the S gene within the receptor binding domain and upstream of the polybasic furin cleavage site. For infection length there were only two mutations (G446S and T547K) significantly associated and both within the spike protein.

**Table 1.**
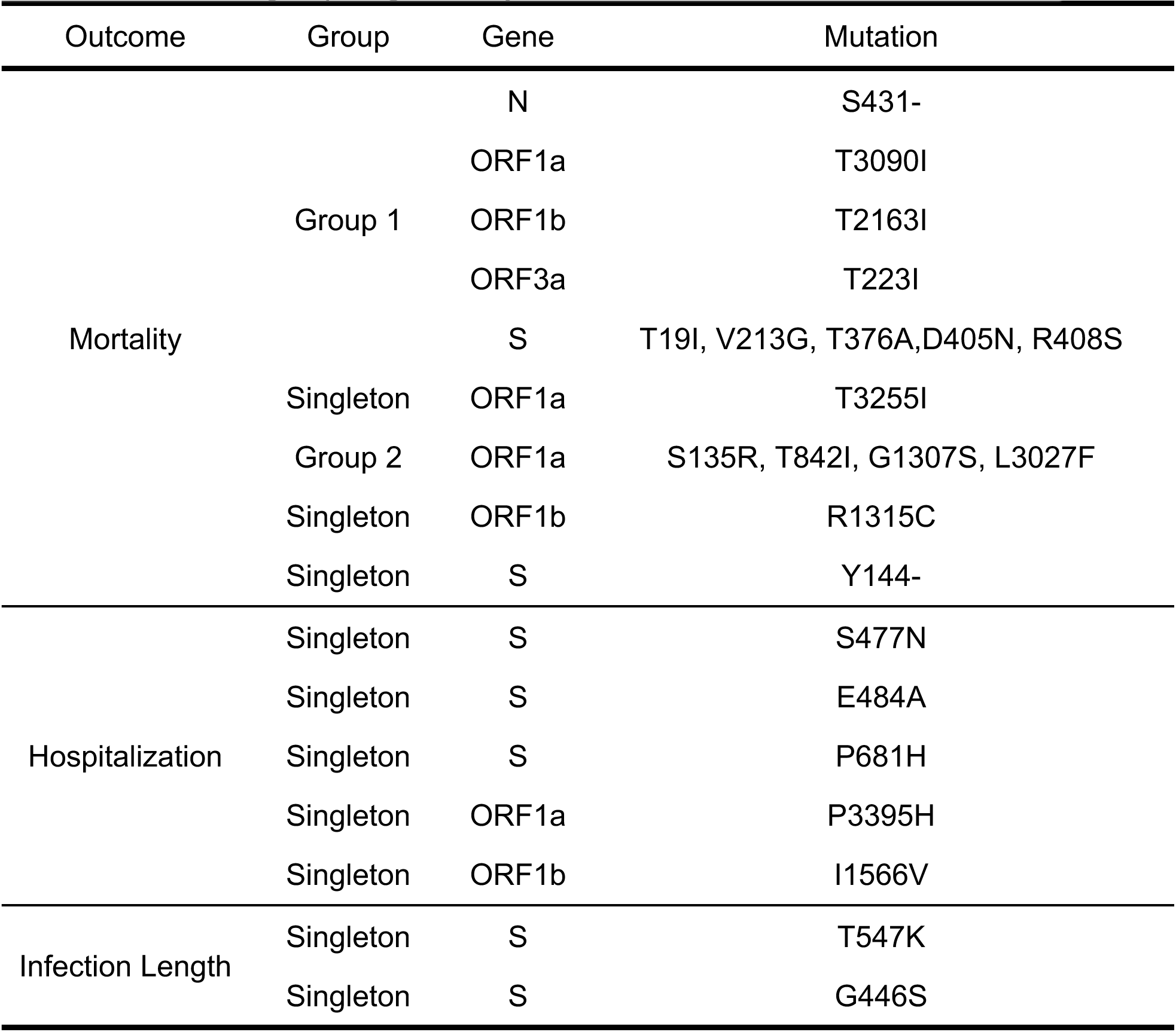
Mutational groups significantly associated with clinical outcomes.

## Discussion

In this study, we demonstrate that systematically enriching pathogen genomic records with sequence-specific patient metadata enhances the analytical power of downstream epidemiological investigations. Among >100,000 articles published in 2023-2024, only ∼0.02% provided feasibly accessible sequence-specific patient metadata. This highlights the limitations faced by scientists performing secondary data analysis as well as deficiencies in data depositing standards, corroborating earlier reports of gaps in genomic data stewardship [7,42]. Through manual curation, we recovered a median of 14 additional metadata variables per sample, including age, sex, comorbidities, treatment, and clinical outcomes, increasing overall metadata completeness by an average of 30%. This enriched dataset facilitated two key findings: infections in immunocompromised patients show increased within-host evolution and incorporating clinical covariates strengthens genotype-phenotype association models. Collectively, we illustrate how richer host information can significantly increase the depth of usable information in sequence databases for precision public health inquiry.

The volume of metadata stored exclusively in unstructured formats, such as article text, figures and supplementary materials, presents significant challenges for scalability and data reuse. Although our manual extraction efforts partially address this gap, the approach is labor intensive and impractical as the number of articles increases. Demographic data like age and sex were the most commonly recovered enriched metadata types (relative to pre-enriched), reflecting standardized sampling strategies and their importance in epidemiologic study design [43]. In contrast, clinical information was rarely recoverable and when present were found in unstructured text or embedded in figures. This scarcity is further compounded by inconsistencies in clinical documentation and privacy regulations enforced by Health Insurance Portability and Accountability Act (HIPAA) or General Data Protection Regulation (GDPR), as a few examples.

To address this gap will require both policy and technical interventions, such as enforcing more metadata deposition, data sharing frameworks that provide as much metadata without sacrificing patient confidentiality, and NLP-based tools for metadata extraction.

Traditional clinical NLP has been used to map narrative text to standardized codes with expert level accuracy, accounting for modifiers including negation, temporal information, and family history [44]. More recently, machine learning pipelines have been developed for the large-scale detection of patient metadata from COVID-19 literature [45]. Incomplete or ambiguous geospatial metadata poses significant challenges for accurate genomic epidemiology, a known issue where host location metadata is inconsistent or missing [17,18]. To address this issue, frameworks have been developed that integrate uncertainty in sampling locations into phylodynamic analysis [19,20]made to develop natural language processing (NLP) based systems designed to automatically extract and refine geospatial data directly from scientific literature [21]. Klein et al., show that fine-tuned models pretrained on biomedical corpora, BiomedBERT, outperforms large language models for classifying metadata containing text. Until development and adoption of NLP-based tools, demographic data will remain more accessible than clinical metadata, limiting population level efforts to identify epidemiological trends.

Longitudinal sequencing from the 34 patients, from our case study, showed that pathogens isolated from immunocompromised individuals contain more amino acid mutations than immunocompetent individuals. Although lineage BA.1 showed a borderline association with elevated rates, we found that overall viral clades had minimal impact on evolutionary dynamics compared to host immune status. Infections in severely immunosuppressed individuals, such as those with hematologic malignancies or organ transplants, can persist for months, effectively allowing the virus to undergo continuous adaptation under reduced immune pressure [46]. This extended evolutionary window not only increases the total number of mutations but also shifts in selective pressures acting on the viral population. We observed that immunocompromised patients harbor significantly more unique amino acid mutations, while unique nucleotide changes showed no significant differences between groups. The higher rates of nonsynonymous mutations suggests that there is likely some positive selection, rather than just neutral drift influencing the viral population [47].

We have shown the utility of integrating detailed clinical metadata with pathogen genomic data for elucidation of mutational dynamics and clinical outcomes. Specifically, we identified 63 mutations with a low probability of occurring by chance and recurred across multiple patients and viral lineages. Of these, the majority of mutations we identified were localized to the spike protein, including many known to be associated with antibody escape (D405N, E484A, G446S, K417N, L452R, N440K, N501Y, N969K, Q493R, and S371F) associated with antibody escape [48–53]. These patterns of convergent mutations are comparable to those noted in other studies of persistent SARS-CoV-2 infections in immunocompromised hosts [54]. Separately, we also recovered two intergenic nucleotide mutations (C241T and A28271T), whose recurrence might suggest some potential regulatory functions that warrant further investigation.

Benefiting from the gains in predictive model performance from incorporating clinical covariates we were able to establish an association between mortality and a pattern of mutations that span both structural and non-structural genes. Specifically, mutations in the S gene (T19I, V213G, T376A, D405N, and R408S) have been shown to enhance the virus’s ability to bind and enter host cells [48,49]. The non-structural mutations (T30901, T2163I, andR1315C) are found in genes known to be involved in both disabling host innate immune signaling and controlling the kinetics of the viral replication [55]. Together these mutations likely have a synergistic effect in combination with host factors (age and comorbidities) that delay or prevent immune response leading to the progression of a more severe and fatal disease.

Most of the mutations we identified as being associated with hospitalization were localized to the binding domain of the spike gene. Specifically, we foundS477N, common among omicron variants and known to increase ACE2 affinity [56], suggesting that enhanced viral entry is a key factor in disease severity. In contrast, the length of infection appears to be influenced by mechanisms of immune invasion. The mutations associated with infection length G446S and T547K, have been linked to altered T-cell recognition [57] and viral fusion phenotypes [58], respectively. While efficiency in viral entry may lead to more severe symptoms, SARS-CoV-2 persistence in the host is mediated by mutations that facilitate immune evasion.

Together, these observations illustrate the potential of integrated metadata-enriched pathogen genomics in facilitating biologically coherent genotype-phenotype relationships.

This study illustrates the significant, yet often unrecognized, advantage of enriching sequence databases with demographic and clinical metadata. Recent audits have shown that approximately 60% of GISAID submissions are missing data [42] or contain errors/ambiguities [7]. These combined lead to misguided genotype-phenotype analyses, compromising the accuracy and reliability of epidemiological studies. Adopting structured and harmonized contextual data standards, such as those developed by the Public Health Alliance for Genomic Epidemiology (PH4GE), would mitigate these issues and maximize the utility of sequencing data across databases and institutions [59].

### Limitations

Our study has several limitations that should be considered when interpreting our findings. First, our literature search was restricted to open-access articles published in 2023 and 2024 with genomic data available in GenBank; this approach omits relevant articles behind paywalls and excludes genomic data that are only available in other databases such as GISAID. This targeted approach led to a dataset that was both geographically and temporally skewed to studies from Asia (69.1%) and from 2022 (76.2%), which limits the broader applicability of our findings to other regions, populations, and phases of the pandemic. Further, despite combining data across four articles, our analysis included only 34 patients. In addition, we did not account for potential differences in access to clinical care and treatment across the four different patient cohorts, which may introduce confounding. Finally, while manual enrichment was effective, the labor-intensive process can introduce inconsistencies in metadata collection, reinforcing the critical need for standardized, structured and accessible deposition of patient metadata with genomic sequences.

## Conclusion

Here we show the importance of enriched host metadata in understanding both viral evolution and clinical dynamics. The explanatory power that comes from linking pathogen genomic data directly with patient characteristics produces a far more comprehensive picture of disease behavior. There have been few large-scale SARS-CoV-2 sequencing studies that have incorporated rich patient metadata, largely because most publicly available genomic sequences contain limited clinical information. Our research directly quantifies the problem and shows how improvement in metadata availability strengthens pathogen genomics. Incorporating enriched metadata has facilitated the identification of genotypic and phenotypic patterns that sequence data alone could not demonstrate. Comprehensive patient metadata (enriched dataset) provides critical insights into the interplay among viral mutations, clinical outcomes, and transmission potential, offering a more complete understanding of viral evolutionary behavior within populations.

## Code Availability

All code used for data processing, analysis and figure generation in this study is publicly available on GitHub at https://github.com/pavia27/Pathogen-Genome-Framework-manuscript/.

## Author Contributions

Conceptualization, M.J.P, G.G., and M.S.; Methodology, M.J.P., K.O., and M.S.; Data Curation, M.J.P. and K.O.; Formal Analysis, M.J.P, K.O., and M.S.; Validation, M.J.P and M.S.; Visualization, M.J.P; Resources, K.O. and G.G.; Writing - Original Draft, M.J.P and M.S.; Writing - Review & Editing, M.J.P., K.S., G.G. and M.S.; Project administration, Supervision, and Funding acquisition, G.G. and M.S.

## Competing Interests

The authors declare no competing interests.

## Data Availability

The metadata enriched genomic dataset analyzed in this study is available in the supplementary file “Supplementary_Data.csv”, it is also dynamically available https://dataviz.hlpgonzalezlab.com/genomic-metadata. Most genomes are publicly available on the NCBI database, and their corresponding accession ids are provided in the “genbank” column. This dataset also includes 167 genomes for which raw sequencing data were obtained from the SRA and assembled (as described in the methods section). The raw SRA accession numbers are also stored in the “genbank” column. Of those SRA assembled genomes 81 have are deposited in the GISAID database and their GISAID accession ids are provided in the “id” column.

## Notes

Funding: Research reported in this publication was supported by the National Institute Of Allergy And Infectious Diseases of the National Institutes of Health under Award Number R01AI164481 to GGH and MS. The content is solely the responsibility of the authors and does not necessarily represent the official views of the National Institutes of Health

### Competing Interest Statement

The authors have declared no competing interest.

### Funding Statement

Research reported in this publication was supported by the National Institute Of
Allergy And Infectious Diseases of the National Institutes of Health under Award Number
R01AI164481 to GGH and MS. The content is solely the responsibility of the authors and does
not necessarily represent the official views of the National Institutes of Health.

### Author Declarations

This study used only openly available human data, all of which were originally located in GenBank (https://www.ncbi.nlm.nih.gov/genbank/) and in the peer-reviewed publications associated with these genome sequences.

