## Supplemental_Material for "A Metadata-Driven Framework for Strengthening Pathogen Genomics Lessons from SARS-CoV-2"

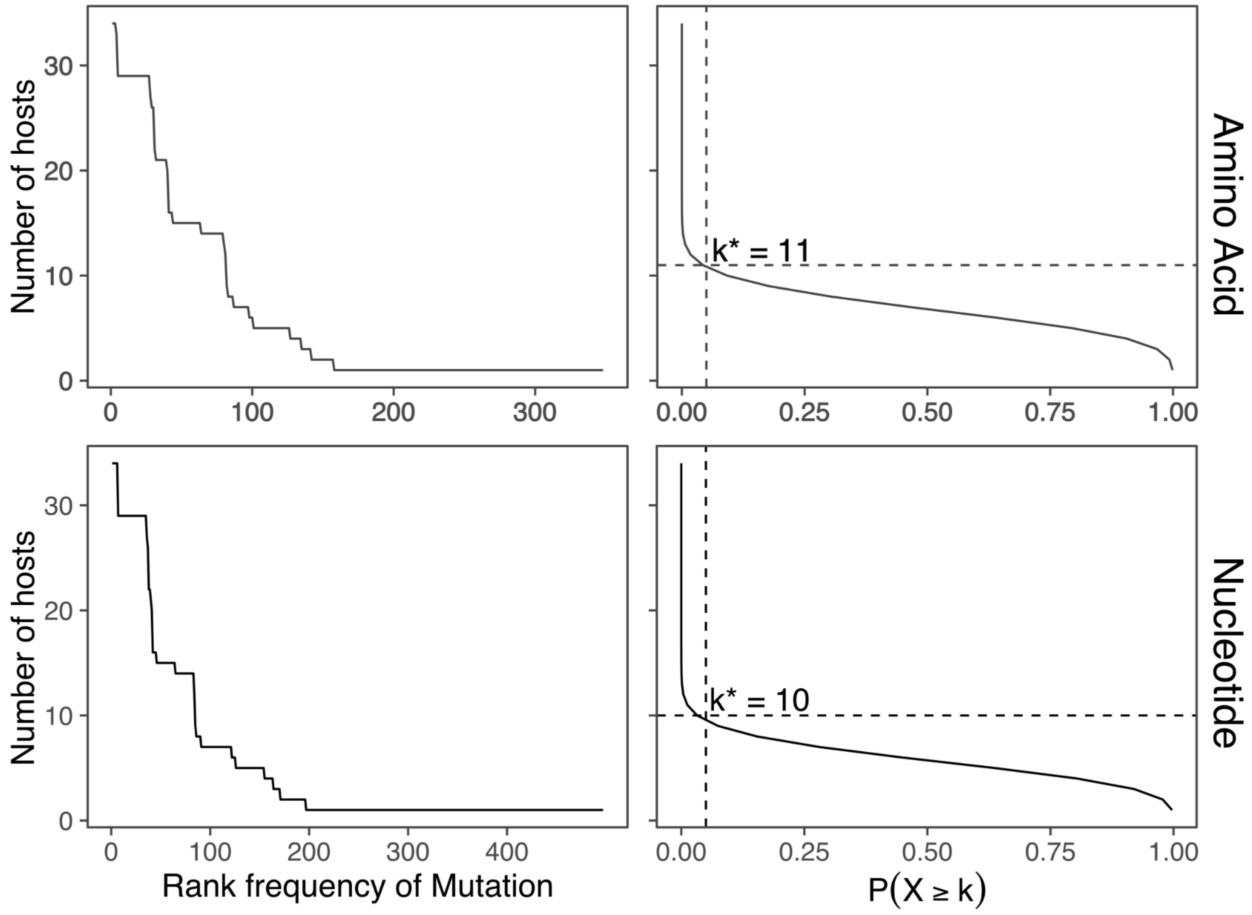


Figure S1.Rank frequency and empirical threshold for non-random mutations. Top panels are from amino acid mutations and bottom panels are from nucleotide mutations. The right panels show the rank frequency distribution of unique mutations. The left panels show the empirical probability under a binomial null model. The dashed vertical (p =0.05) and horizontal (minimum patient count exceeding P =0.05) lines indicate the significance cut-off.

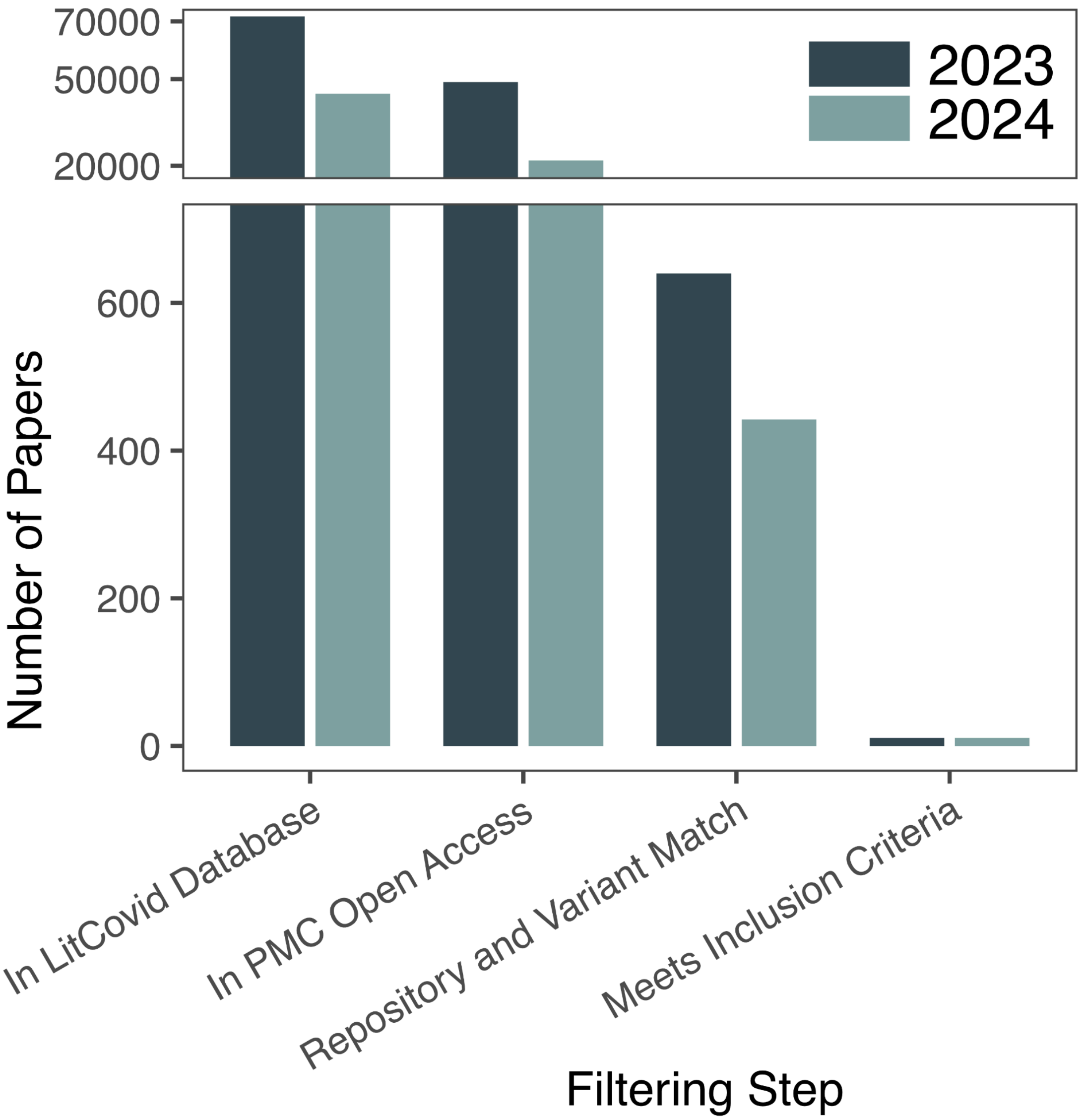


Figure S2. Number of article records retained at each stage of the filtering process. For each year counts are shown for (1) all articles in LitCovid, (2) open access full texts that were successfully downloaded, (3) articles with matching regular expressions for both sequence database and VOC/VUM, (4) articles included after manual review that met inclusion criteria.

Table S1. Characteristics of the 21 SARS-CoV-2 sequencing studies that were included in metadata enrichment.

| Article ID | Time Span | Country | Genomes | Patients | PMID |
|---|---|---|---|---|---|
| Chen 2024 | 1/22 - 4/22 | China | 72 | 72 | 38316363 |
| Gonzalez_Reiche_2023 | 1/22 - 5/22 | USA | 13 | 4 | 37270625 |
| Igari_2024 | 8/22 - 8/22 | Japan | 3 | 1 | 38684722 |
| Jin_2023 | 6/22 - 7/22 | Japan | 10 | 9 | 37744926 |
| Jony_2024 | 12/23 - 1/24 | Bangladesh | 14 | 14 | 38651907 |
| Liu_2023 | 1/22 - 6/22 | Taiwan | 13 | 13 | 37789031 |
| Liu_2024 | 1/22 - 5/23 | China, Singapore | 572 | 572 | 38191416 |
| Manuto_2024 | 12/21 - 3/22 | Italy | 37 | 22 | 38543811 |
| Misra_2023 | 1/22 - 11/22 | India | 47 | 47 | 37684328 |
| Pavia_2024 | 3/22 - 2/23 | Italy | 9 | 8 | 38804179 |
| Penas_Utrilla_2023 | 1/22 - 7/22 | Spain | 6 | 6 | 37460980 |
| Perez_Florido_2023 | 2/22 - 5/22 | Spain | 29 | 29 | 36768752 |
| Prost_2023 | 2/22 - 12/22 | France | 138 | 138 | 37544942 |
| Sayama_2024 | 2/22 - 2/22 | Japan | 1 | 1 | 38890805 |
| Selvavinayagam_2023 | 9/22 - 1/23 | India | 89 | 89 | 38076717 |
| Singh_2024 | 5/22 - 8/22 | India | 5 | 5 | 38724525 |
| Taboada_2023 | 4/22 - 8/22 | Mexico | 6913 | 6913 | 38112714 |
| Tahsin_2024 | 12/23 - 1/24 | Bangladesh | 17 | 17 | 38656213 |
| Tsai_2024 | 8/22 - 10/22 | Taiwan | 2 | 2 | 39388868 |
| Ulhuq_2023 | 1/22 - 3/22 | Scotland | 56 | 56 | 37083576 |
| Zhao_2024 | 3/22 - 3/22 | China | 61 | 61 | 39493536 |

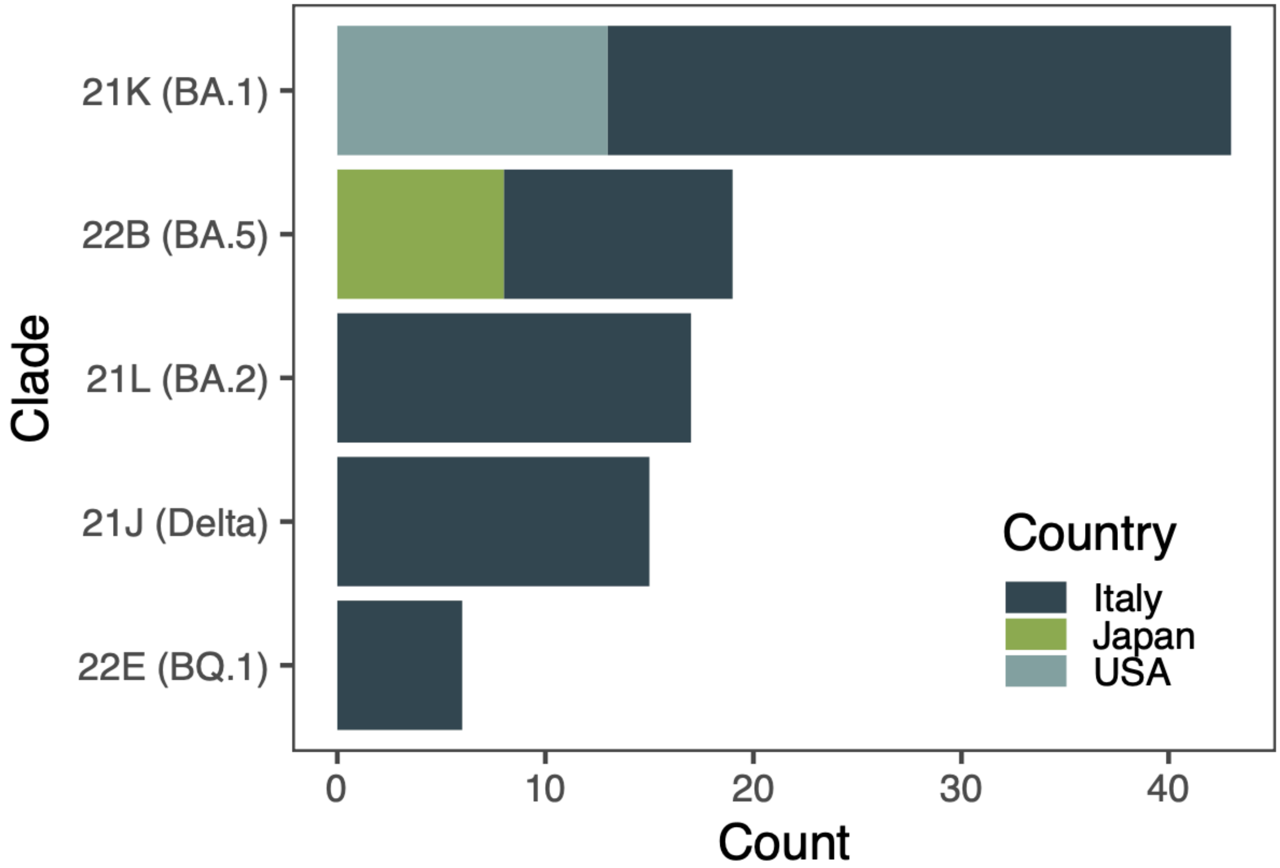


Figure S3. Distribution of SARS-CoV-2 lineages by country.
